# Inferring the number of COVID-19 cases from recently reported deaths

**DOI:** 10.1101/2020.03.10.20033761

**Authors:** Thibaut Jombart, Kevin van Zandvoort, Timothy W Russell, Christopher I Jarvis, Amy Gimma, Sam Abbott, Sam Clifford, Sebastian Funk, Hamish Gibbs, Yang Liu, Carl A. B. Pearson, Nikos I Bosse, Centre for the Mathematical Modelling of Infectious Diseases COVID-19 Working Group, Rosalind M Eggo, Adam J Kucharski, W John Edmunds

## Abstract

We estimate the number of COVID-19 cases from newly reported deaths in a population without previous reports. Our results suggest that by the time a single death occurs, hundreds to thousands of cases are likely to be present in that population. This suggests containment via contact tracing will be challenging at this point, and other response strategies should be considered. Our approach is implemented in a publicly available, user-friendly, online tool.

---

As the coronavirus-2019 (COVID-19, (1)) epidemic continues to spread worldwide, there is mounting pressure to assess the scale of epidemics in newly affected countries as rapidly as possible. We introduce a method for estimating cases from recently reported COVID-19 deaths. Results suggest that by the time the first deaths have been reported, there may be hundreds to thousands of cases in the affected population. We provide epidemic size estimates for several countries, and a user-friendly, web-based tool that implements our model.

## Using deaths to infer cases

COVID-19 deaths start to be notified in countries where few or no cases had previously been reported (2). Given the non-specific symptoms (3), and the high rate of mild disease (4), a COVID-19 epidemic may go unnoticed in a new location until the first severe cases or deaths are reported (5). Available estimates of the case fatality ratio, *i*.*e*. the proportion of cases that are fatal (CFR, (6,7)), can be used to estimate the number of cases who would have shown symptoms at the same time as the fatal cases. We developed a model to use CFR alongside other epidemiological factors underpinning disease transmission to infer the likely number of cases in a population from newly reported deaths.

Our approach involves two steps: first, reconstruct historic cases by assuming non-fatal cases are all undetected, and, second, model epidemic growth from these cases until the present day to estimate the likely number of current cases. We account for uncertainty in the epidemiological processes by using stochastic simulations for estimation of relevant quantities.

Two pieces of information are needed to reconstruct past cases: the number of cases for each reported death, and their dates of symptom onset. Intuitively, the CFR provides some information on the number of cases, as it represents the expected number of deaths per case, so that CFR^−1^ corresponds to the expected number of cases per death. In practice, the number of cases until the first reported death can be drawn from a Geometric distribution with an event probability equal to the CFR. Note that while our approach could in theory use different CFR for each case (to account for different risk groups), our current implementation uses the same CFR for all cases in a simulation. Dates of symptom onset are simulated from the distribution of the time from onset to death, modelled as a discretised Gamma distribution with a mean of 15 days and a standard deviation of 6.9 days (8).

Once past cases are reconstructed, we use a branching process model for forecasting new cases (9,10). This model combines data on the reproduction number (*R*) and serial interval distribution to simulate new cases ‘*y*_*t*_’ on day ‘*t*’ from a Poisson distribution:

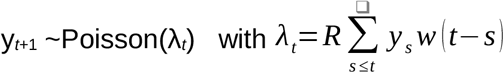

where w(.) is the probability mass function of the serial interval distribution. More details on this simulation model can be found in Jombart *et al*. (10). Optionally, this model can also incorporate heterogeneity in transmissibility using a Negative Binomial distribution instead of Poisson. The serial interval distribution was characterised as a discretised Lognormal distribution with mean 4.7 days and standard deviation 2.9 days (11). We assume that past cases caused secondary transmissions independently (i.e. are not ancestral to each other), so that simulated cases for each death can be added. This assumption is most likely to be met when reported deaths are close in time. As the time between reported deaths increases, past cases may come from the same epidemic trajectory rather than separate, additive ones, in which case our method would overpredict epidemic size.

Further details on model design and parameters values are provided in Supplementary Material. Our approach is implemented in the R software (12) and publicly available as R scripts (see Supporting Information) as well as in a user-friendly, interactive web-interface available at: https://cmmid.github.io/visualisations/inferring-covid19-cases-from-deaths.

## How many cases for a single death?

We first used our model to assess likely epidemic sizes when an initial COVID-19 death is reported in a new location. We ran simulations for a range of plausible values of *R* (1.5, 2 and 3) and CFR (1%, 2%, 3% and 10%), assuming a single death on the 1st March 2020 (7). 25,000 epidemic trajectories were simulated for each parameter combination. Simulations for an ‘average severity’ scenario (7) with *R* = 2 and CFR = 2% show that by the time a death has occured, hundreds to thousands of cases may have been generated in the affected population (Figure 1). Results vary widely across other parameter settings, and amongst simulations from a given setting (Table 1), with higher *R* and lower CFR leading to higher estimates of the numbers of cases. However, a majority of settings give similar results to our ‘average’ scenario, suggesting that a single death is likely to reflect several hundreds of cases. Results were qualitatively unchanged when incorporating heterogeneity in the model using recent estimates (13), but prediction intervals were wider (Supplementary Material).

**TABLE 1:**
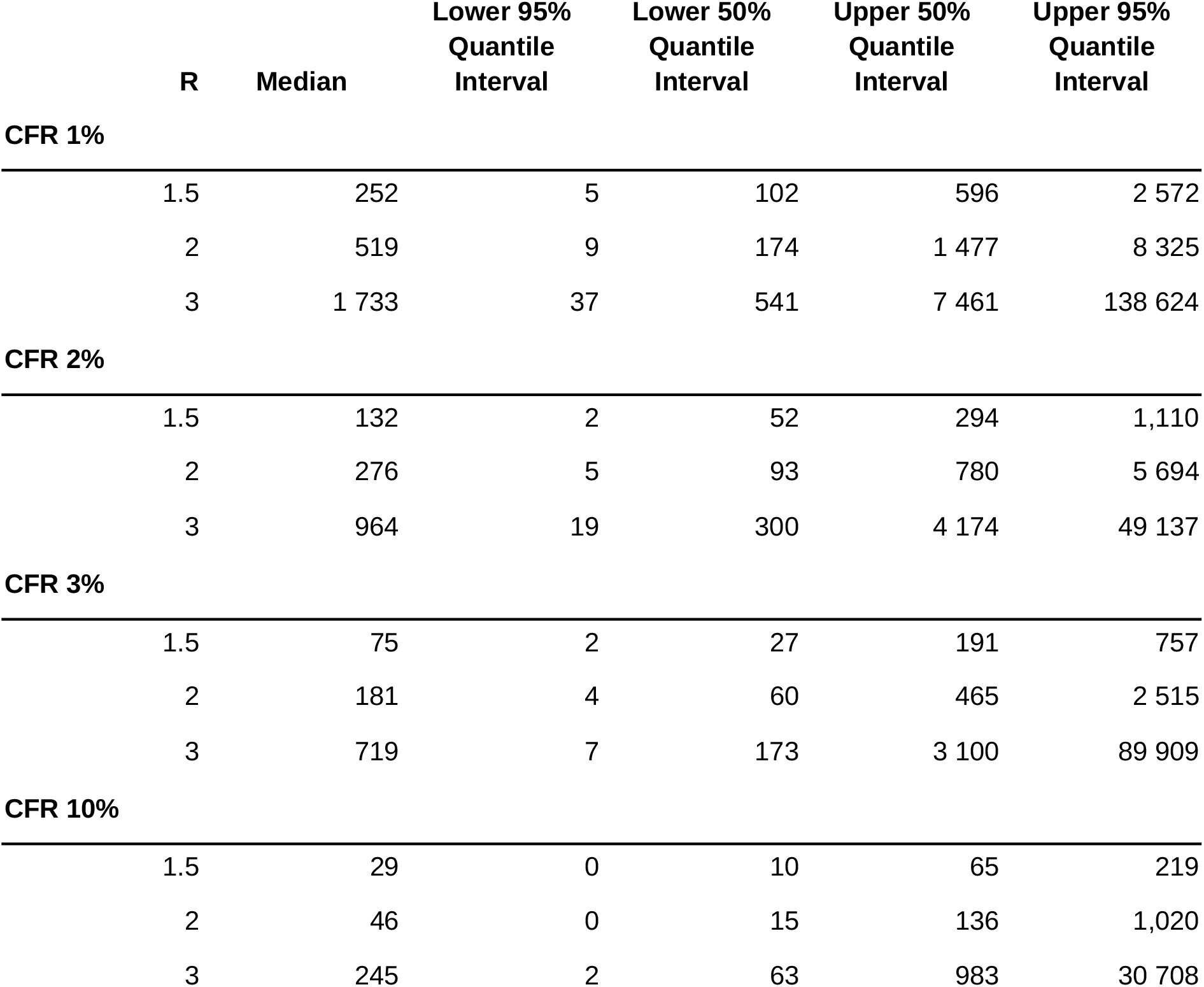
inferred number of cases for a single death. Inferred number of cases after detection of a single death under different values of the reproduction number, and case fatality ratio. We estimate the number of expected cases in the population at the day the death occurred, and present median, 50%, and 95% estimates of the quantile interval.

**Figure 1.**
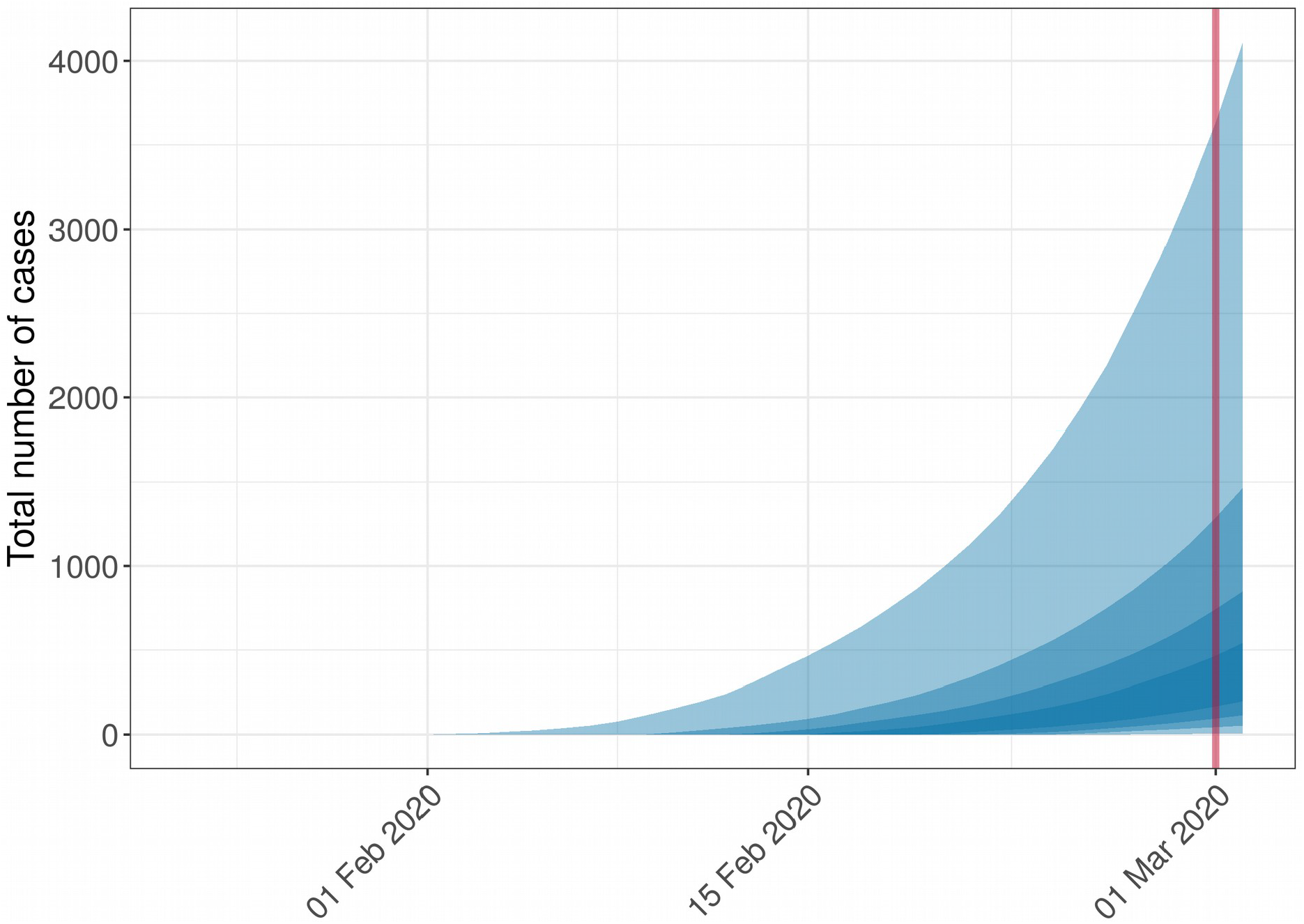
Example of simulated epidemic trajectories from a single death. This figure shows results of 200 simulations using a CFR of 2% and *R* of 2 based on an hypothetical situation where a single death occurred on the 1st March 2020, represented by the red line. Ribbons of different shades represent, from the lightest to the darkest, the 95%, 75%, 50% and 25% quantile intervals.

## Recently affected countries

We applied our approach to three countries which recently reported their first COVID-19 deaths (Spain, Italy, and France), using the same range of parameters as in the single-death analysis. In order to compare predictions to cases actually reported in these countries, projections were run until 4th March. Overall, predictions from the model using the baseline scenario (*R* = 2, CRF = 2%) were in line with reported epidemic sizes (Table 2). Results from other scenarios are presented in the Supplementary Material. Actual numbers of reported cases fell within the 50% quantile intervals of simulations in all three countries Italy (median: 1 294 ; QI_50%_: [390 ; 3 034]; reported: 2 037), France (median: 592 ; QI_50%_: [177 ; 1 705]; reported: 190) and Spain, (median: 202 ; QI_50%_: [95 ; 823]; reported 202).

**TABLE 2:** Inferred number of cases for several countries assuming CFR of 2% and R of 2. All values are presented for the 4th of March 2020 for different countries. We present the predicted case counts as their median, 50%, and 95% estimates of the quantile interval. * First suspected death due to within country transmission.

| Country | Date of first death* | Initial deaths | Reported cases | Median | Lower 95% Quantile Interval | Lower 50% Quantile Interval | Upper 50% Quantile Interval | Upper 95% Quantile Interval |
| --- | --- | --- | --- | --- | --- | --- | --- | --- |
| Spain | 4th March | 1 | 202 | 263 | 8 | 95 | 823 | 7 829 |
| Italy | 26th Feb | 1 | 2 037 | 1 294 | 33 | 390 | 3 034 | 19 487 |
| France | 21st Feb | 1 | 190 | 592 | 10 | 177 | 1 705 | 7 501 |

## Discussion

Several limitations need to be considered when applying our method. First, our approach only applies to the deaths of patients who have become symptomatic in the location considered, which should usually be the case in places where traveller screening is in place. We also assume constant transmissibility (*R*) over time, which implies that behaviour change and control measures have not taken place yet, and that there is no depletion of susceptible individuals. Consequently, our method should only be used in the early stages of a new epidemic, where these assumptions are reasonable. Similarly, the assumption that each death reflects independent, additive epidemic trajectories is most likely to hold true early on, when reported deaths are close in time (e.g. no more than a week apart). Used on deaths spanning longer time periods, our approach is likely to overestimate epidemic sizes.

Contact tracing has been shown to be an efficient control measure when imported cases can be detected early on (14), in addition to permitting the estimation of key epidemiological parameters (11). When the first cases reported in a new location are mostly deaths, however, our results suggest that the underlying size of the epidemic would make control via contact tracing extremely challenging. In such situations, efforts focusing on social distancing measures such as school closures and self-isolation may be more likely to mitigate epidemic spread.

## Data Availability

Our model is implemented in a publicly-available web-application (see link provided).

https://cmmid.github.io/visualisations/inferring-covid19-cases-from-deaths

## ACKNOWLEDGEMENTS

The named authors (TJ, SA, AG, CIJ,, TWR, KvZ, SC, SF, HG, YL, CP, NIB, RME, AJK, WJE) had the following sources of funding:

TJ, CIJ and AG receive funding from the Global Challenges Research Fund (GCRF) project ‘RECAP’ managed through RCUK and ESRC (ES/P010873/1). TJ receives funding from the UK Public Health Rapid Support Team funded by the United Kingdom Department of Health and Social Care. TJ receives funding from the National Institute for Health Research - Health Protection Research Unit for Modelling Methodology. SA and SF were funded by the Wellcome Trust (grant number: 210758/Z/18/Z), TWR and AJK were funded by the Wellcome Trust (grant number: 206250/Z/17/Z), SC was funded by the Wellcome Trust (grant number: 208812/Z/17/Z), and RME was funded by HDR UK (grant number: MR/S003975/1). KvZ is supported by the Elrha’s Research for Health in Humanitarian Crises (R2HC) Programme. The R2HC programme is funded by the UK Government (DFID), the Wellcome Trust, and the UK National Institute for Health Research (NIHR). HG is funded by the Department of Health and Social Care using UK Aid funding and is managed by the NIHR. YL receives funding from the National Institute for Health (grant number: 16/137/109) and from the Bill and Melinda Gates Foundation (grant number: INV-003174).

The UK Public Health Rapid Support Team is funded by UK aid from the Department of Health and Social Care and is jointly run by Public Health England and the London School of Hygiene & Tropical Medicine. The University of Oxford and King’s College London are academic partners. The views expressed in this publication are those of the authors and not necessarily those of the National Health Service, the National Institute for Health Research or the Department of Health and Social Care.

## AUTHORS CONTRIBUTIONS

TJ developed the model and the app, and wrote the first draft of the manuscript.

WJE, TJ, TR, CIJ, AK, SC, RE, CABP conceived the method.

AG, CIJ, SA, SF, KvZ. contributed code.

TR, YL, HG, AG, CIJ contributed data.

CIJ, SA, KvZ contributed analyses.

SA, SC, AG, CABP, NB, CIJ reviewed code.

TJ, CIJ, SA, AG, RE, AK, JE, KvZ, NB, SC contributed to the manuscript.

CMMID COVID-19 Working Group gave input on the method, contributed data and provided elements of discussion. The following authors were part of the Centre for Mathematical Modelling of Infectious Disease 2019-nCoV working group: Mark Jit, Charlie Diamond, Fiona Sun, Billy J Quilty, Kiesha Prem, Nicholas Davies, Stefan Flasche, Alicia Rosello, James D Munday, Petra Klepac, Joel Hellewell. Each contributed in processing, cleaning and interpretation of data, interpreted findings, contributed to the manuscript, and approved the work for publication.

All authors read and approved the final version of the manuscript.

